# Genome Profiling of Actionable Cancer Targets (NYU LG-PACT) for Clinical Patient Molecular Diagnostics and Treatment

**DOI:** 10.64898/2026.08.27.26361341

**Authors:** Yiying Yang, Varshini Vasudevaraja, Jonathan Serrano, Hussein Mohamed, Stephen Kelly, George Jour, Tatyana Gindin, Kyung Park, Derek Jones, Xiaojun Feng, Jared Pinnell, Samantha Mclennan, Man Ying Tin, Aristotelis Tsirigos, Matija Snuderl, Kazimierz O. Wrzeszczynski

## Abstract

Next-generation sequencing (NGS) for the detection of somatic variants has become the method of choice in a variety of molecular oncology fields and in the clinic. Its use ranges from sequencing entire tumor genomes and transcriptomes to targeted clinical diagnostic gene panels. The NYU Langone Genome PACT (Profiling of Actionable Cancer Targets, LG-PACT) assay is a qualitative *in vitro* diagnostic test that uses targeted next generation sequencing (NGS) of formalin-fixed paraffin-embedded (FFPE) tumor tissue matched with normal specimens from patients to detect gene alterations in a targeted panel covering 606 genes and the TERT promoter. Indications for testing are cancer (solid tumors and hematological malignancies) where a mutational profile from multiple genes would be informative for disease stratification, prognosis, or treatment options including targeted therapies and eligibility for clinical trials. The test is intended to provide information on somatic mutations including point mutations, small insertions/deletions (indels), and copy number aberrations for diagnostic and treatment decisions. LG-PACT is a United States Food and Drug Administration (FDA) cleared diagnostic test (510K: K202304). The clinical interpretation of sequencing data of molecular tumor markers from NGS encompasses automated variant calling tools with human interpretation. This final mostly manual review of data step is intensive, involving highly trained scientists, encompassing literature review, interpretation and clinical tier classification by pathologists, who then provide a complete molecular diagnostic report to the treating oncologists. We provide analysis of 1339 clinical genomic profiles from 31 different cancers and their subtypes, comprising of central nervous system (CNS) 792 (59%) cases (incl. meningioma, glioma and glioblastoma), with 267 (20%) cases predominantly of lung, pancreatic and colorectal and 280 of others (21%). Here, we present the technical challenges of validating an NGS oncological diagnostic targeted assay for clinical grade accuracy and sensitivity for patient care. We show how copy number alterations provide a more comprehensive description of the tumors genomic profile. We then outline the utility of targeted panel sequencing based on certified pathologist selection of reportable variants for our current patient cohort. Where analysis of variant detection has led to 49.4% (661/1339) of our clinical tumor samples containing mutations in known therapy targeted genes, 35.6% (477/1339) with mutation detected in other genes, and 15% (201/1339) cases being negative.

## Introduction

Over the past decade, genomic characterization of somatic alterations in cancer has been conducted both retrospectively to survey the genomic landscape of human malignancies^1–3^ and prospectively, to inform patient care strategies^4–6^. Next Generation Sequencing (NGS)-based diagnostic assays have become indispensable tools within hospital pathology departments for diagnosing and managing both somatic and inherited disease^7–12^. Analysis of these expansive datasets reveals that the utility of precision medicine, driven by large-scale genomic profiling, is intricately dependent on the specific variant, cancer type, and patient context^13–16^. While it is undeniable that sequencing hundreds of genes or even whole genomes in malignant neoplasms, coupled with the discovery and regulatory approval of NGS-based biomarkers, has enabled oncologists to offer individualized treatment options that were unimaginable a decade ago^17,18^, the implementation of genomic profiling in clinical oncology remains nuanced^19^ and continued efforts to expand the clinical genome is ongoing. Currently, most genomic profiling in hospital settings is performed using targeted NGS panels of varying sizes^20–24^. This approach is driven by multiple factors, including platform accessibility and cost, rapid turnaround times, biomarker-based eligibility for clinical trials, FDA approval status, and reimbursement considerations^25,26^.

At NYU Langone Health, the molecular pathology department has developed a FDA-approved 606-gene panel plus the TERT promoter region for use in both solid tumors and hematologic malignancies that supports clinical decision-making. This panel enables comprehensive mutational profiling across multiple genes, providing insights into disease stratification, prognosis, and therapeutic options, including targeted therapies and clinical trial eligibility.

In this report, we first outline the technical challenges involved in validating an NGS-based targeted oncology assay to meet clinical-grade standards for accuracy and sensitivity. We then present real world data from the genomic profiles of our hospital’s patient cohort sequenced between 2022 and June 2024 using this targeted panel. Finally, we analyze pathologist-guided clinically reported variants and discuss the current capabilities of precision oncology, as enabled by targeted NGS panels in guiding patient management.

## Methods

### NYU LG-PACT Assay

The NYU Langone Genome PACT (LG-PACT) is a matched tumor-normal custom targeted panel assay analyzing a set of 606 gene targets and TERT promoter region (**Supplementary Table 1**) validated for both solid tumors and hematological malignancies. The LG-PACT is a pan-cancer solid tumor panel for detection of single nucleotide variants (SNV), small insertions and deletions (indel) and focal copy number variants (CNV) including amplifications, hemi- and homozygous deletions. The NYU LG-PACT sequencing panel contains all genes classified as Therapeutic Level 1 by OncoKB^TM^ ^27^.

### DNA Sequencing

DNA from FFPE, fresh tissue, bone marrow, or peripheral blood is extracted using the Maxwell (Promega Corporation, Madison, WI) automated nucleic acid extraction system as per manufacturers guidelines for each specimen type. An estimated minimum of 10% tumor content as assessed by a pathologist is required for sequencing. DNA sequencing libraries were prepared with the Kapa HyperPlus Kit (Roche, Basel, Switzerland). Extracted genomic DNA (250ng, with minimum of 100ng) undergoes enzymatic fragmentation, end repair and A-tailing, adapter (Integrated DNA Technologies, Coralville, Iowa) ligation, post-ligation bead cleanup, and library amplification. Each sample’s libraries are then pooled and undergo hybridization capture with a custom set of IDT XGen lockdown probes targeted to designated genes in our NYU LG-PACT gene panel, and undergo Streptavidin bead capture and clean up, with final PCR enrichment. Post capture library sizes were measured with 4200 TapeStation (Agilent, Santa Clara, Ca) quantified by Qubit High Sensitivity Kit (ThermoFisher Scientific, Waltham, MA). DNA sequencing is then performed as a 150bp paired-end reads on the Illumina NextSeq 550 platform (Illumina, San Diego, CA) targeting a validated minimum of 300X for tumor and normal with the average tumor coverage as 1074X (std. dev. 347X) and 701X (std. dev. 138X) for normal in our cohort.

### Bioinformatics Analysis

LG-PACT assay involves use of various Off the Shelf (OTS) software tools for the identification of somatic alterations SNVs, INDELs (<=35bp) from the patient’s tumor DNA compared to matching normal DNA (Mutect2 ^28^ and LoFreqSomatic ^29^). NYU LG-PACT identifies and reports copy number (CN) aberrations per individual gene for amplifications plus either homozygous and hemizygous deletions using the CNVkit software package 24 ^30^. Gene amplifications are identified when gene copy number is greater than 4 while hemizygous loss (CN = 1) are clinically reported only when associated with a SNV or indel in the same gene. Complete homozygous loss defined as both copies of a gene deleted (CN = 0), and is always reported. Hypermutation status was determined based on variant burden per megabase of sequencing panel (>10 mutations/Mb). Given the LG-PACT panel covers approximately 2.1 Mb, cases with ≥20 variants were classified as hypermutant. The entire informatics pipeline is integrated together for both the pre and post processing of sequencing data. The entire bioinformatics workflow is outlined below and includes the following steps (**Figure 1**). Pre-processing: GATK BWA-MEM alignment of FASTQ files, marking and removing duplicates, applying GATK best practices ^31^, and performing quality control (QC). Alignment is to the GRCh37/hg19 reference human genome. Somatic/Germline Variant Calling and Annotation: Realigned and recalibrated tumor-normal BAM files undergo variant calling, copy number variation calling, and processing of germline samples. Post-processing Annotation and Custom Filtering: VEP annotated somatic alterations (using MANE Select and OncoKB^TM^ transcripts) are then subjected to various custom filtering steps to make sure only variants within 95% confidence intervals are pushed to manual review by pathologists. Variants are reviewed using the Philips Intellispace Precision Medicine (ISPM) software system and variants selected by pathologists were exported from Philips ISPM and included in this study.

**Figure 1:**
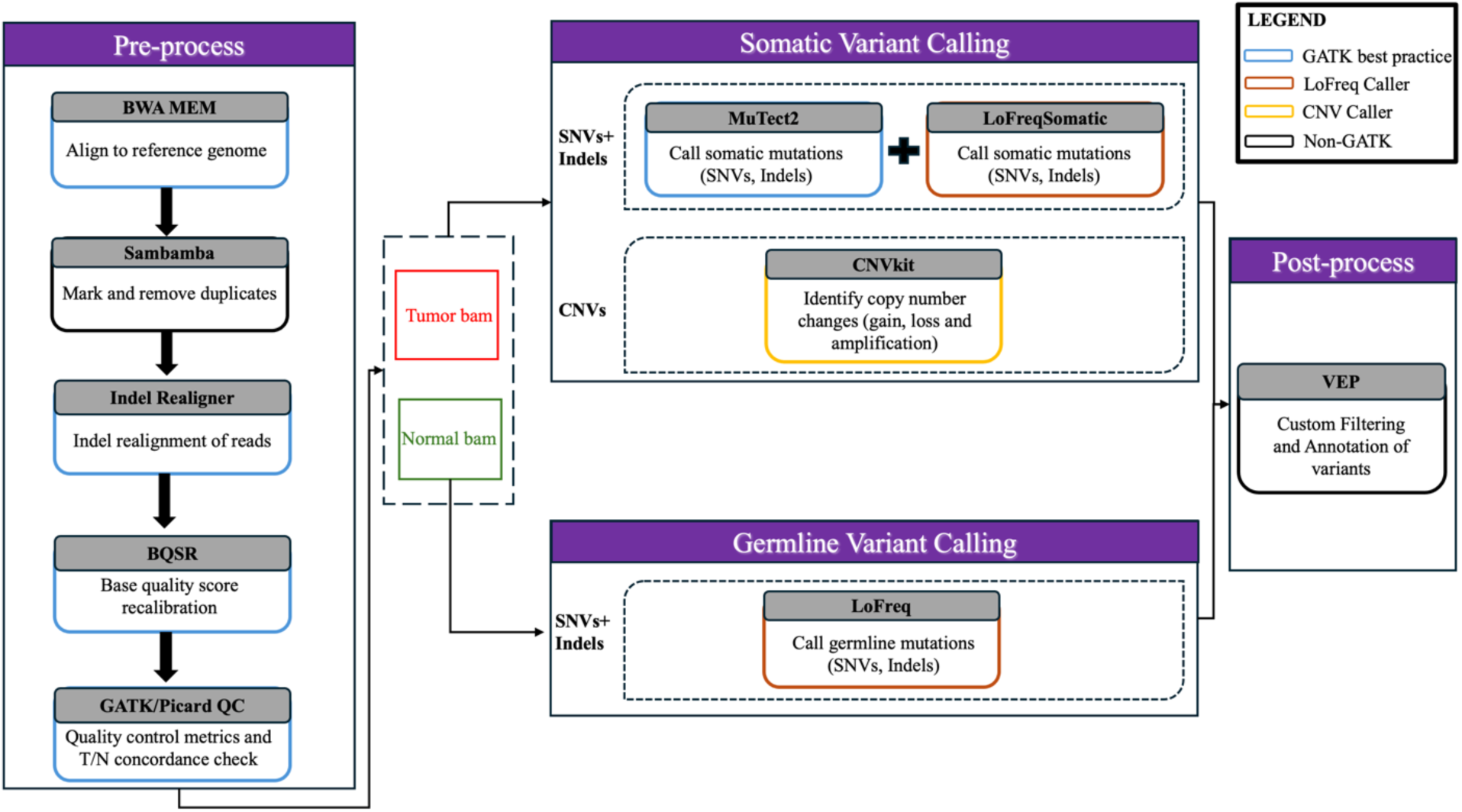
Overview of NYU LG-PACT assay workflow. The analysis of tumor-normal samples involves three main steps: **1.** Pre-processing: Includes aligning the FASTQ files, marking and removing duplicates, applying GATK best practices, and performing quality control (QC). **2.** Variant Calling: Realigned and recalibrated tumor-normal BAM files undergo variant calling, copy number variation (CNV) calling, and processing of germline samples. **3.** Post-processing: Annotation and Custom Filtering: All alterations are annotated, and custom filtering based on validated clinical performance thresholds is applied.

### LG-PACT bioinformatics workflow framework and implementation in clinical setting

To support the NYU LG-PACT targeted NGS assay, we implemented a robust, scalable, and modular pipeline using the Nextflow workflow management system ^32^. Each analytical stage of the assay (**Figure 1**), from raw data quality control through somatic variant calling and annotation, is encapsulated within version-controlled Singularity containers. This containerization strategy ensures consistent software environments and reproducible execution across both high-performance computing (HPC) clusters and cloud-based platforms. The result of our entire pipeline workflow which is processed using 4 nodes on a HPC is approximately 27-30 hours upon completion of sequencing. The Nextflow framework was selected based on its strong support for modular pipeline design, containerization, automatic logging, error handling, and seamless integration with clinical compliance requirements (CAP/CLIA). Variants are reviewed using the Philips ISPM and final reports are subsequently pushed to the EPIC electronic health record system, facilitating integration with clinical workflows and supporting precision oncology efforts.

### Patient cohort

Our patient cohort includes 1339 patients who completed the NYU LG-PACT assay between 2022 and June 2024 (**Table 1**). The median patient age was 60.6 years old with slightly higher female prevalence (54.4%). Median turnaround time is 17 days from receipt of paired specimens to final report sign out and import into an EPIC patient portal. Patients were diagnosed with a diverse range of tumors spanning 31 types and subtypes. Cancer assignment per tumor sample (case) was based on the diagnosis for interpretation as designated by the ordering physician. The most common diagnoses within our health system at this time included meningioma (17.6%), glioma (11.4%), and lung cancer (11.1%), followed by pituitary adenoma (9.6%), and schwannoma (8.4%). The cohort also included 27 cases (2%) of cancers of unknown primary diagnosis. Self-reported ethnicity data from the NYU Langone Health electronic healthcare record (EHR) system EPIC revealed that only 27% of the cohort represents the ethnic diversity of the city our hospital is based in and 11% of the cohort with no ethnicity data reported. For those with reported ethnicity, 62% are white, 10.6% are Asian, 9.2% Black or African American, 5.1% Hispanic or Latino or Spanish, and 1.8% reported more than one ethnicity. Cancer type distributions differed between cases with and without ethnicity reported (**Supplementary Figure 1A**). Among cases with major ethnicity categories reported (**Supplementary Figure 1B**), the most common cancer type varies. Meningioma cases are predominated among White, Black, and Hispanic patients (e.g., 23.4% within Black patient group), whereas lung cancer was most frequent among Asian patients (19.1%). Pituitary adenoma contributed a larger share of cases among Black and Asian patients than among White patients in this cohort.

**Table 1.**
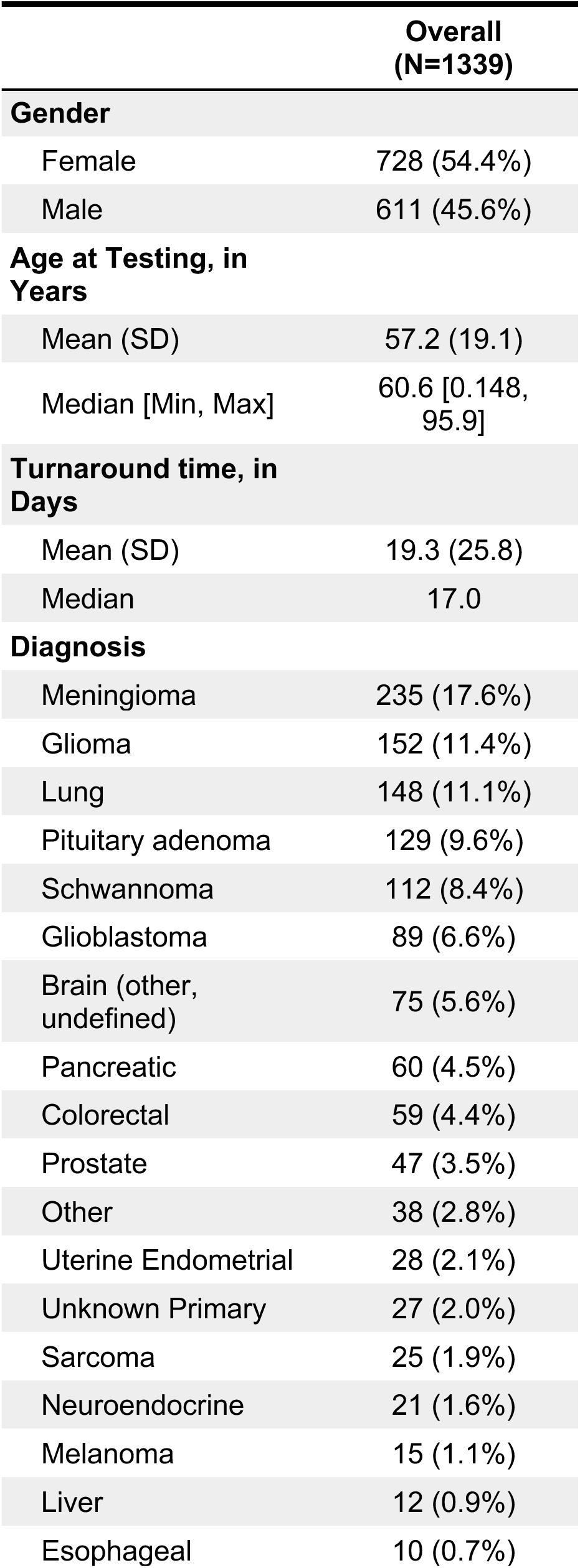
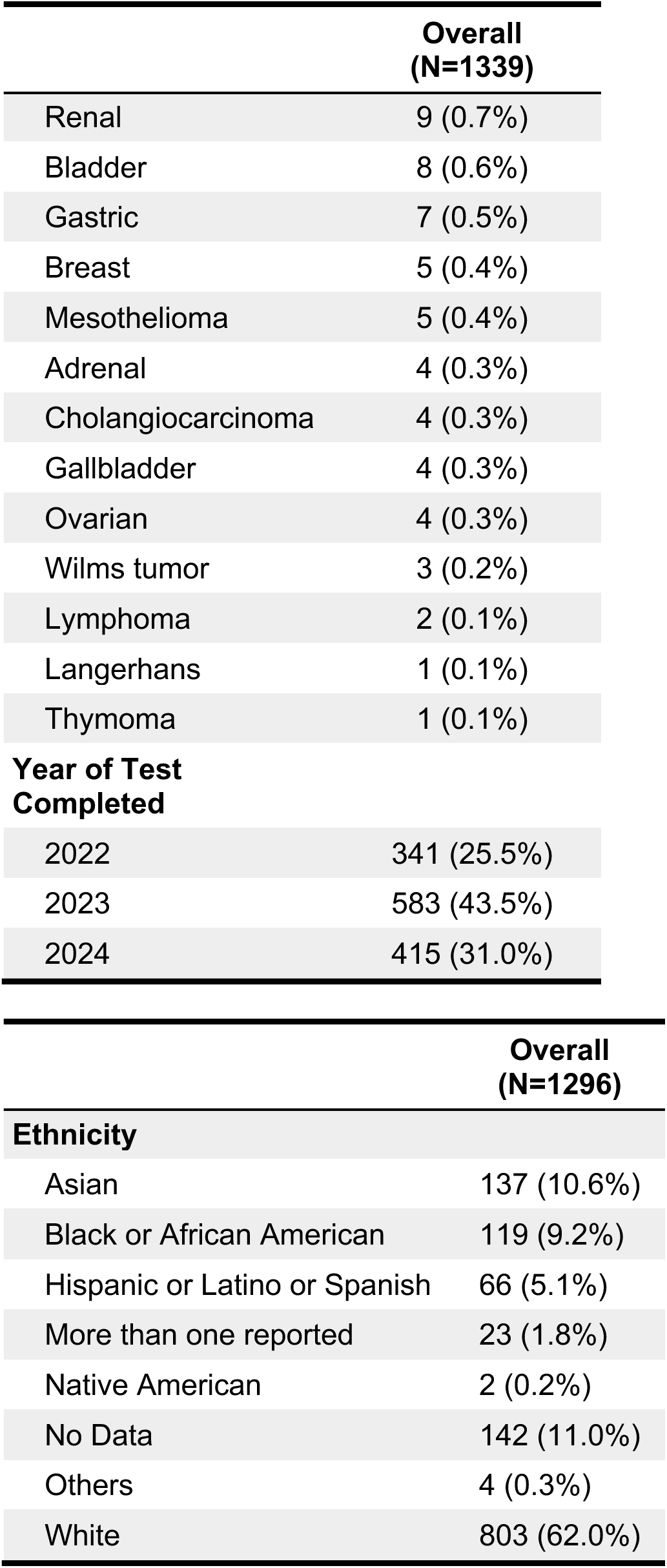
Cohort clinical characteristics and diagnostic overview of 1339 cases profiled by NYU LG-PACT between 2022 and June 2024 (1296 unique patients). NYU LG-PACT cohort summary for gender, age, turnaround time, clinical diagnosis and ethnicity of cases processed at our institution between January 2022 to June 2024. A total of 1339 cases from 1296 unique patients with 42 patients undergoing testing more than once. Turnaround time was calculated as number of days between test completion date and specimen received date. Age was calculated as years between the patient’s date of birth and the specimen received date. Ethnicity data were obtained from self-reported ethnicity fields in NYU Langone Health EHR (EPIC). Ethnicity categories were based on options available at the time of reporting. For patients who selected more than one category, they were classified under “More than one reported”.

## Results

### Variant calling validation and performance

FDA and New York State validation of variant calling of the NYU LG-PACT oncology panel was performed by assaying 306 orthogonally identified variants in 145 samples from 10 cancer tissue types and 44 genes containing 270 SNVs, 18 deletions and 9 insertions (**Supplementary Table 2).** Variant allele frequency (VAF) was significantly correlated (Pearson correlation = 0.97) with a slope close to one when compared to the previously orthogonally reported variants (**Supplementary Figure 2**). The establishment of confidence interval thresholds for sequencing coverage and variant allele frequency limit of detection were established using previously applied power calculations ^20,33^ to minimize false-negative results. In summary, approximately 100X coverage is determined to be necessary to detect mutations present at 10% VAF with 0.95 power and 500X coverage is necessary for 95% confidence for detection of a variant with 5% frequency (based on a 2% error rate or minimum Q20 Phred score). We finalized >300X average sequencing read coverage which enables a detection of mutations present at 6%VAF with 0.95 power.

We then applied these variant calling validation thresholds and determined variant allele frequency (VAF) and read coverage of variants identified in all clinical cases (n=1339) profiled between 2022 and June 2024 by our assay using two variant callers Mutect2 ^28^ and LoFreqSomatic ^29^ (**Figure 2, Supplementary Figure 3**) and with filtering of all variants for 54 OncoKB^TM^ Therapeutic Level 1 genes (**Figure 2, Supplementary Table 1**). Overall, somatic variant calling of single nucleotide variants (SNVs) by NYU LG-PACT gene panel from 1339 tumor-normal pairs identified a total of 8024 SNVs (an average of 6 per tumor-normal pair) and 2508 somatic insertions and deletions (indels) (an average of 1.9 per tumor-normal pair) (**Table 2**) within thresholds of >=200X and >=5% VAF. Variants common to both variant callers MuTect2 and LoFreqSomatic were 69% for SNVs but only 27% for indels depicting the continued challenge of highly accurate indel calling and the diversity of the two indel calling algorithms. Variants detected by both variant callers are more prominent in the range of our clinically validated thresholds and less prominent below validated thresholds (**Figure 2, Supplementary Figure 3**). A total of 1002 (13%) SNV and 97 (4%) indels called by both variant callers in Level 1 genes were identified.

**Figure 2:**
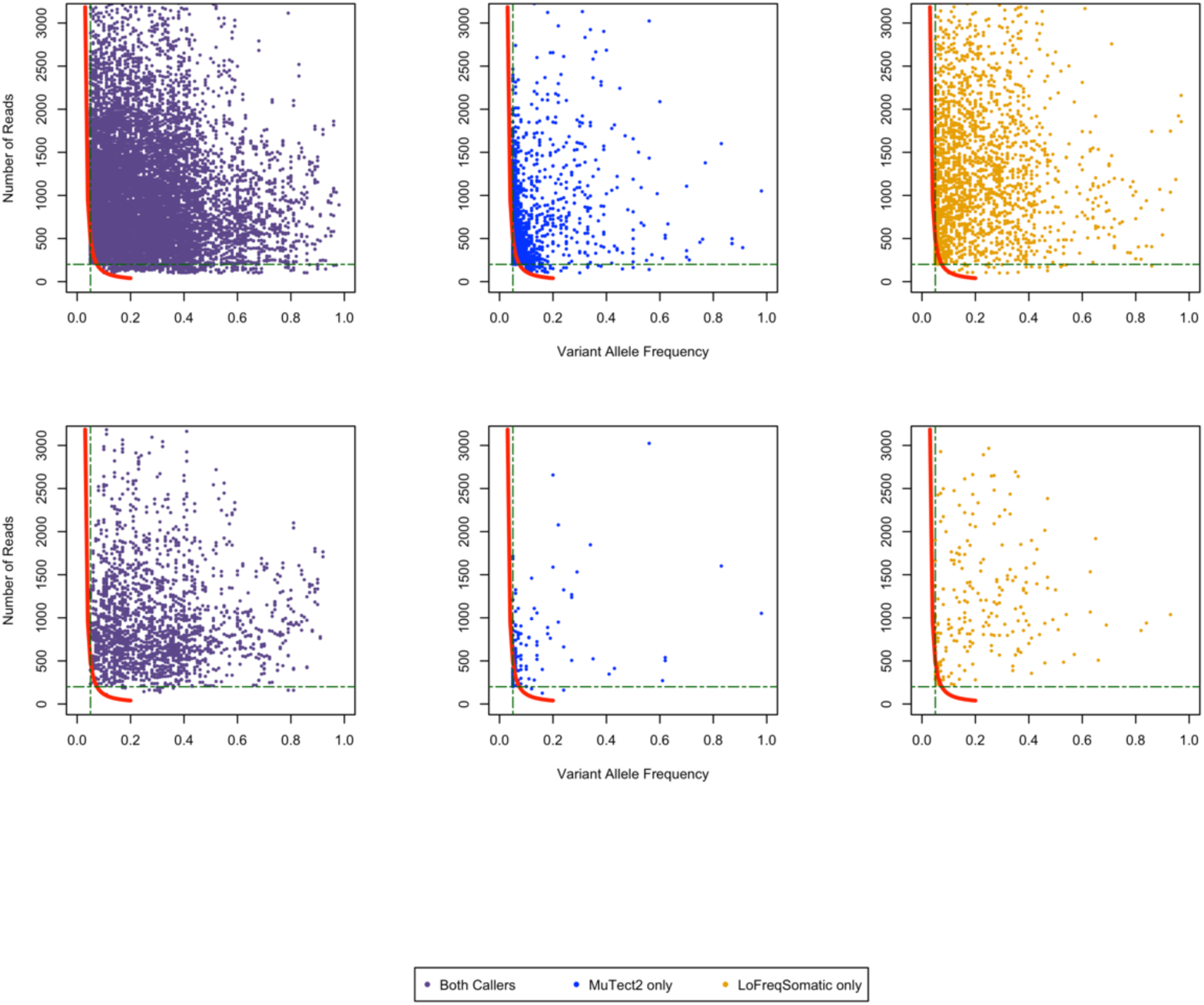
Empirical determination of variant calling performance based on validation filter criteria to detect variants at high-confidence level. SNVs and Indels were identified empirically with MuTect2 and LoFreqSomatic variant callers. We present read count and VAF of variants identified in all clinical cases (n=1339) profiled between 2022 and June 2024 by our assay using two callers (top) and with filtering of all variants for OncoKB^TM^ Tier 1 genes (bottom). Red line – 95% confidence interval (Cheng D., et al. JMDI 2015:17), Blue points – sample variants (both SNV and Indels) called by both MuTect2 and LoFreqSomatic, Gray points – sample variants unique to the one caller. Green dotted lines – horizontal line represents coverage 200x threshold, vertical line represents VAF thresholds at 5%.

**Table 2.** SNVs and Indels identified based on validation criteria. Total counts of SNVs and Indels identified by NYU LG-PACT assay from 2022 – June 2024 (n=1339). Two-caller (variant common to both LofreqSomatic and MuTect2) and single caller variants are shows for all data and in a subset of 54 genes classified as OncoKB^TM^ Therapeutic Level 1.

| Variant Caller | SNVs | INDELs |
| --- | --- | --- |
| **All Data** |  |  |
| MuTect2 | 769 | 1314 |
| LoFreqSomatic | 1721 | 520 |
| Two-Caller | 5534 | 674 |
| **Level 1 Genes** |  |  |
| MuTect2 | 68 | 81 |
| LoFreqSomatic | 179 | 75 |
| Two-Caller | 1002 | 97 |

### SNV/Indel and Copy Number Variant Analysis

Reviewing the somatic mutational profile of our patient cohort, within all 1339 cases, a significant portion of the patients (84.7%, 1134/1339) had at least one mutation reported by pathologists. The distribution of reported mutations per case showed that most cases contained only a few mutations as captured by our panel. The median number of SNV/Indels per sample reported is 4 (**Figure 3A**) (with the average number of SNV/Indels per sample for all 1339 cases reported by NYU pathologists being 6.8). Among the cases analyzed (**Figure 3A**), 62 cases met a hypermutation threshold of ≥20 variants, with 3 of these exhibiting an extreme mutation phenotype (>100 variants). Out of the 606 genes and TERT promoter targeted by NYU LG-PACT panel, 581 (95.7%) genes harbored at least one reported SNV or Indel, while 40 genes were reported only once in our entire cohort (**Supplementary Table 3)** To further illustrate the distribution of aberrations across genes, we examined the relationship between the total number of variants reported per gene and the number of unique variants identified per gene (**Figure 3B**). This analysis revealed that while some genes, such as TP53 and NF2, were reported with both high frequency and broad variant diversity, others, including KRAS, IDH1, and of course the targeted TERT promoter were also frequently reported but with a limited set of hotspot mutations. Prominent genes mutated in 827 cases in our dataset show complex genomic profile containing SNV and indels including frameshift, nonsense and upstream promoter mutations. Overall, from the NYU LG-PACT patient positive cohort, the most commonly reported gene is TP53 (37%), followed by TERT promoter (26%), NF2 (19%), KRAS (16%), and IDH1 (11%) (**Supplementary Figure 4**). When examining beyond the genes and looking more closely at frequent point (or hotspot) mutations, our panel captured the TERT promoter mutations (c.-124C>T and c.-146C>T), and IDH1 (R132H) hotspot mutation which aligns with the significant representation of brain tumor samples in our cohort. The most frequent variants captured by our panel span 19 genes, and 25 of the 50 variants having been designated as oncogenic Tier 1 hotspots by our pathologists (**Supplementary Figure 5**). Indels represented only 10% of frequently reported SNV/Indel mutations with the EGFR exon 19 deletion E746_A750 being the most represented.

**Figure 3.**
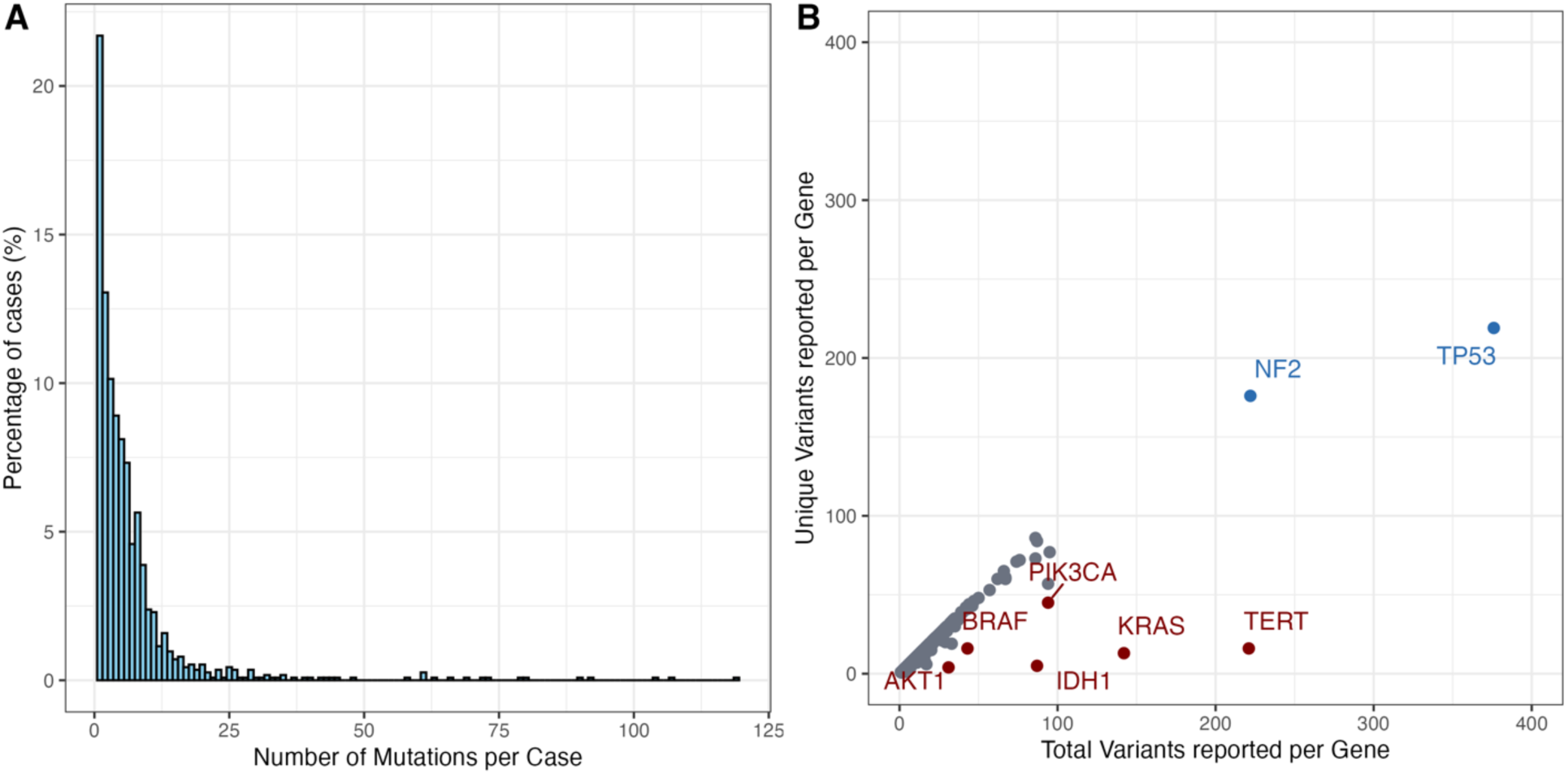
Distribution of mutations (SNV/Indel) per case. **A)** Distribution of the number of SNV/indel variant selected per case (histogram; bin width = 1). The y-axis shows the percentage of cases in each count category (100 × cases in bin / total number of cases). **B)** Per gene, total variant reports (x-axis) versus number of distinct amino-acid–level variant (y-axis), for genes with at least one selected SNV/indel. Highly reported genes with more than 50% variants being unique (blue; >200 total reports), genes with ≥30 reports and ≤50% distinct variants diversity (maroon), and all other genes (gray). Selected genes in the blue and maroon are labeled.

When examining copy number variations, CNVs were detected in 31.9% of cases in our cohort (427/1339), with a median of 1 aberration per CNV-positive sample. We then evaluated whether tumor content is correlated with the likelihood of detecting CNVs. Cases with tumor content ≥50% show more copy number aberrations reported, which is consistent with our validated limit of detection threshold. (**Supplementary Figure 6**). In our clinical reporting criteria, only amplification, hemizygous loss and complete homozygous loss are reported. Gene amplifications are identified when gene copy number is greater than 4 while hemizygous loss (CN = 1) is clinically reported only when associated with a SNV or indel in the same gene. Complete homozygous loss defined as both copies of a gene deleted (CN = 0) is always reported. In total, 204 cases with amplification, 211 cases with hemizygous loss, and 117 cases with homozygous loss were identified. Frequently observed amplifications and deletions as detected by NYU LG-PACT consist of known cancer genes often observed containing structural aberrations across much of the entire genome (**Figure 4, Supplementary Figure 7**). Reportable CNV’s were detected in 63% of cases (837/1339) with average number of copy number aberrations 2.01 per positive sample.

**Figure 4.**
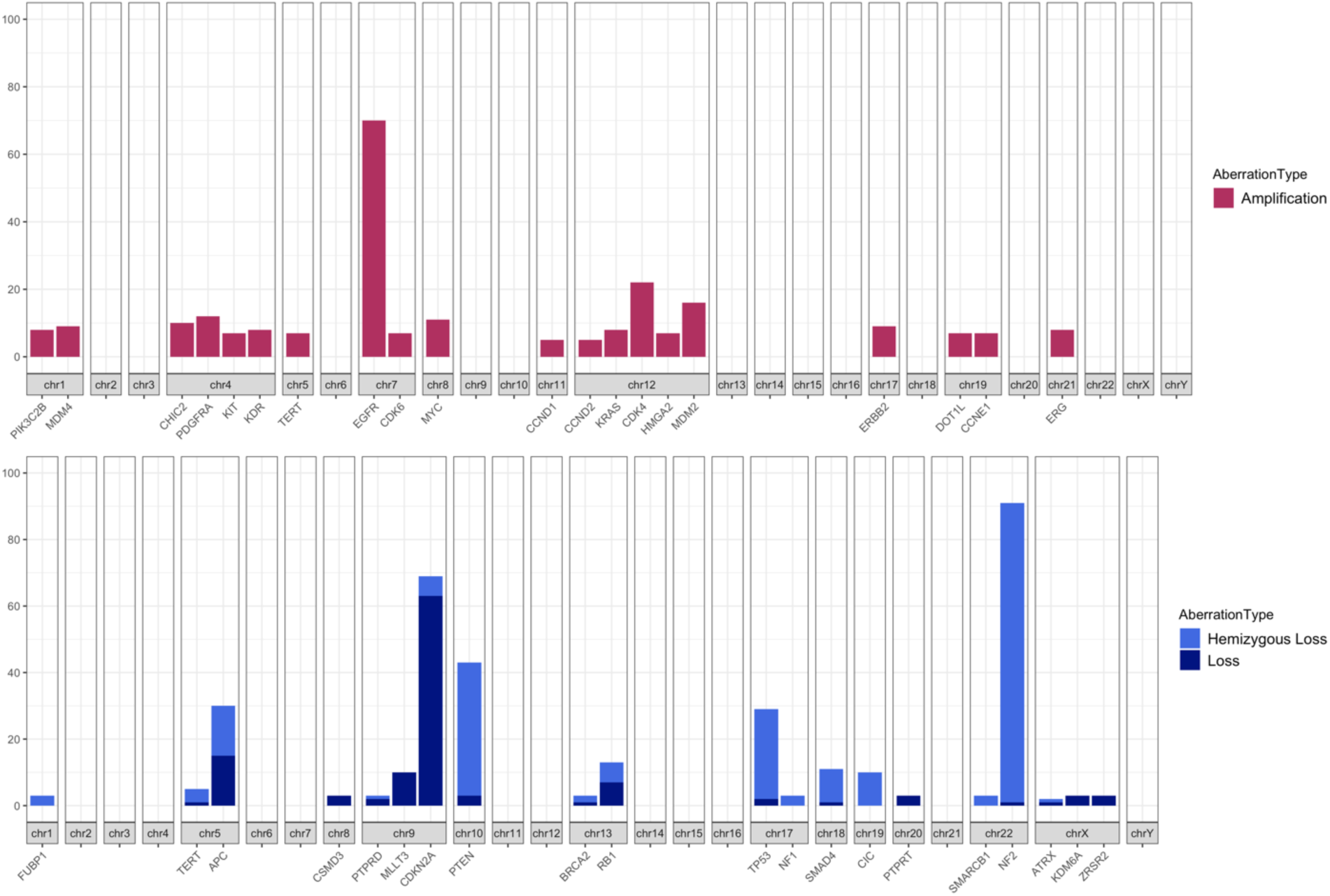
Genomic overview of predominantly reported amplifications and deletions. Percentage of cases with gene-level copy-number aberrations mapped by aberration types and chromosome. The upper figure shows amplifications (copy number > 4). The lower figure shows hemizygous loss (copy number = 1) and deletion (copy number = 0) Genes are arranged according to their corresponding chromosomal locations.

### Clinical Cancer Profiling

We summarize the clinical significance of genes identified per cancer type using OncoKB™(v4.26) Therapeutic Level 1 gene annotations^34^ (Mapping between the OncoKB™ Levels of Evidence and the AMP/ASCO/CAP Consensus Recommendation. https://www.oncokb.org/therapeutic-levels#version=AAC) calculating the number of patient samples containing Level 1 gene mutations per cancer type. Among the most frequently sequenced cancer types in our cohort, lung cancer samples contain the most Level 1 variants reported (**Figure 5**). Certain brain cancers such as glioma and GBM, as well as colorectal and pancreatic cancers, also have reported mutations in Level 1 genes.

**Figure 5.**
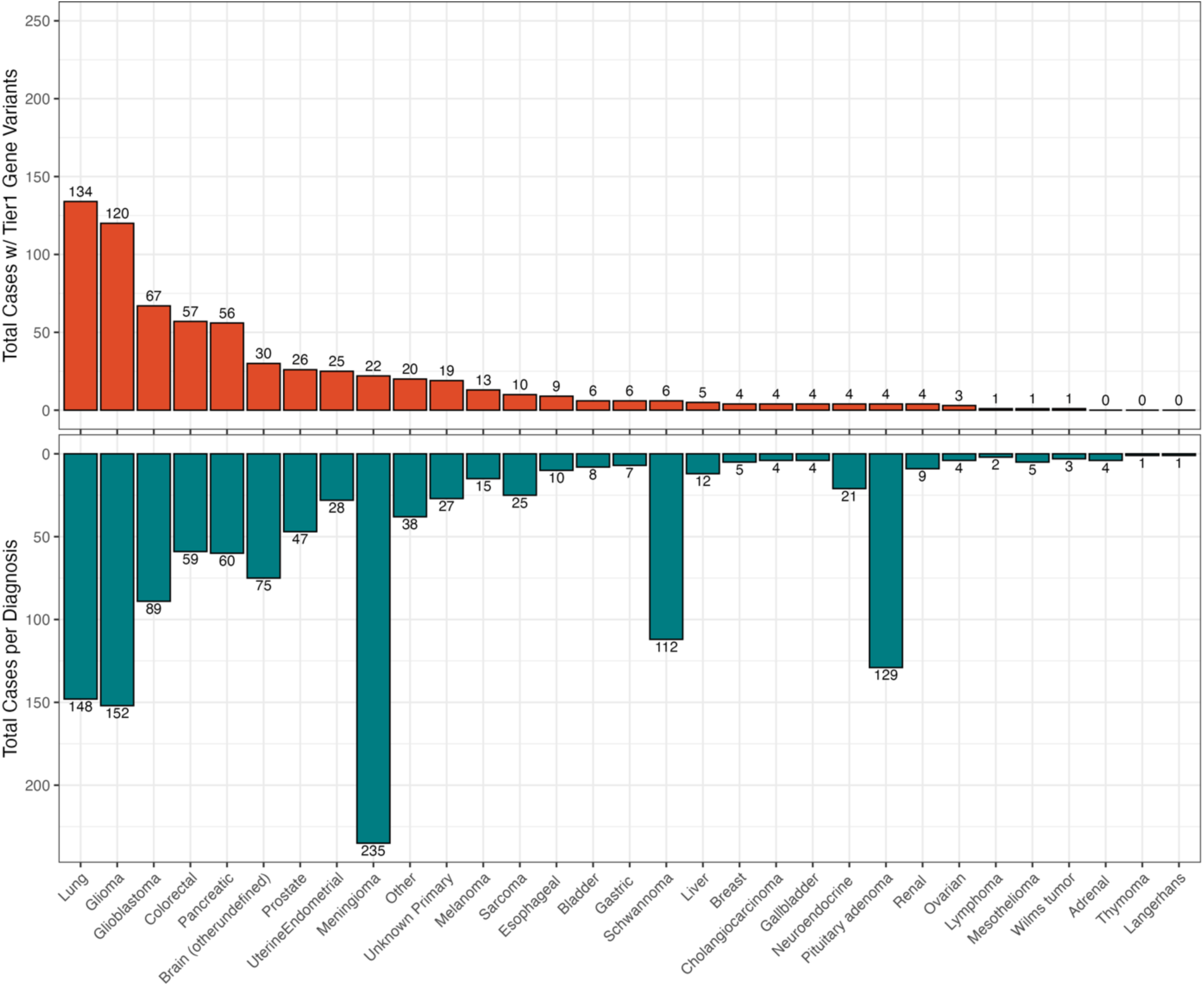
Cancer Mutation Burden from NYU LG-PACT for Targeted Genes. Cancer utility of the NYU LG-PACT assay is illustrated by the total number of cases per cancer type with a mutation in a Therapeutic Level 1 gene (top) and the total number of cases per cancer type in the NYU LG-PACT assay (bottom). Diagnoses are sorted by descending Level 1-positive case count. Number of cases are shown on top of each bar.

Overall, 86% of our patient cohort had either an identified SNV or CNV, with only 14% of cases being completely negative with no aberrations reported. The majority of sequenced patients contained an SNV or Indel (85% of the entire cohort, 1138/1339), with 54% (661/1138) of those cases involving a variant in a therapeutic Level 1 gene. A genomic profile containing both CNV and SNV aberrations was seen in 31% (416/1339) of patient samples, demonstrating the utility of our cancer panel in providing complementary genomic profiling of both single nucleotide aberrations and structural variation (**Figure 6**).

**Figure 6.**
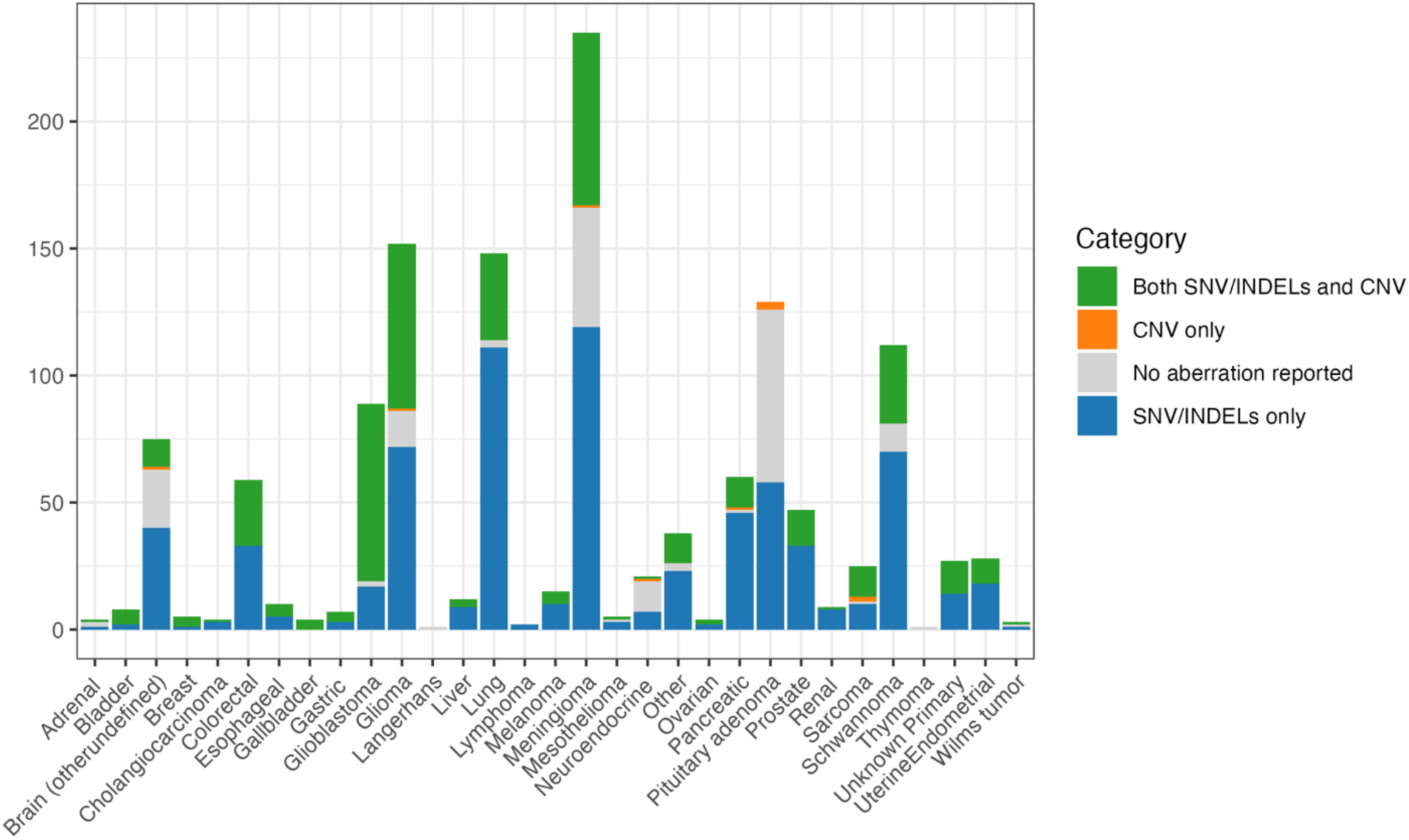
Case-level classification of somatic findings by cancer type in the NYU LG-PACT cohort. Stacked bars show the number of cases per diagnosis, colored by whether the case had SNV/indels only with no reportable CNV (blue), CNV only with no reportable SNV/indels (orange), both SNV/indels and CNV (green), or no somatic SNV/indel or CNV reported (gray). Cancer types are shown in alphabetical order on the x-axis; the y-axis is the number of cases.

Our panel assay identified 96.6% (57/59) of colorectal patients as having known relevant clinical biomarkers in at least one Level 1 gene, including APC, TP53, and KRAS. Lung (90.54%), pancreatic (93.33%), esophageal (90%), and uterine endometrial (89.29%) cancers also show a significant percentage of cases harboring mutations in Level 1 genes (**Table 3**). Notably, all 4 gallbladder cases, though a small cohort, had at least one mutation reported in Level 1 genes.

**Table 3.** NYU LG-PACT panel utility per cancer. For each cancer type, the frequency of cases identified by NYU LG-PACT with somatic variant is shown. ‘Level 1 genes’ refers to the percentage of cases within that cancer type with at least one SNV/Indel variant reported in an OncoKB™ Therapeutic Level 1 gene. ‘Other genes’ represents cases with variants reported in genes that are outside the OncoKB™ level 1 gene list. ‘Negative’ indicates cases with no reported variants.

| **Diagnosis** | **Level 1 genes** | **Top 3 Level 1 genes** | **Other genes** | **Top 3 Other genes** | **Negative** |
| --- | --- | --- | --- | --- | --- |
| Gallbladder (4) | 100% | TP53, ALK, ATM | 100% | AKAP9, ERBB3, KNL1 | 0% |
| Cholangiocarcinoma (4) | 100% | BRIP1, FGFR2, NF1 | 100% | SEP9, CDH2, CDKN2A | 0% |
| Colorectal (59) | 97% | TP53, KRAS, PIK3CA | 100% | APC, SYNE1, SMAD4 | 0% |
| Pancreatic (60) | 94% | KRAS, TP53, ATM | 85% | SMAD4, CDKN2A, ARID1A | 4% |
| Lung (148) | 91% | TP53, EGFR, KRAS | 96% | LRP1B, CSMD3, KEAP1 | 3% |
| Esophageal (10) | 90% | TP53, PIK3CA, ABL1 | 100% | CSMD3, NFE2L2, CDKN2A | 0% |
| UterineEndometrial (28) | 90% | PIK3CA, TP53, ALK | 97% | ARID1A, PTEN, DST | 0% |
| Melanoma (15) | 87% | NRAS, BRAF, ALK | 87% | TERT, CACNA1E, DCC | 0% |
| Gastric (7) | 86% | KIT, TP53, BARD1 | 86% | ARFGEF2, DST, FBXW7 | 0% |
| Breast (5) | 80% | TP53, ALK, ATR | 100% | KMT2C, RNF213, ANKRD26 | 0% |
| Glioma (152) | 79% | IDH1, TP53, NF1 | 88% | TERT, ATRX, CIC | 10% |
| Glioblastoma (89) | 76% | TP53, EGFR, PIK3CA | 98% | TERT, PTEN, LZTR1 | 3% |
| Bladder (8) | 75% | TP53, ATR, KRAS | 100% | TERT, KMT2D, MYH9 | 0% |
| Ovarian (4) | 75% | TP53, ATR, ALK | 100% | CSMD3, ZRSR2, ADGRA2 | 0% |
| Unknown Primary (27) | 71% | TP53, KRAS, PIK3CA | 100% | ARID1A, KMT2C, ASPM | 0% |
| Prostate (47) | 56% | TP53, PIK3CA, ATM | 98% | FOXA1, APC, SPOP | 0% |
| Other (38) | 53% | TP53, BRAF, KRAS | 87% | KMT2D, CSMD3, LRP1B | 8% |
| Lymphoma (2) | 50% | BRAF | 100% | ADGRL3, B2M, DST | 0% |
| Renal (9) | 45% | TP53, ATM, BRIP1 | 100% | PBRM1, VHL, ACVR2A | 0% |
| Liver (12) | 42% | TP53, ATM, NRAS | 100% | TERT, CTNNB1, ARID1A | 0% |
| Brain (otherundefined) (75) | 40% | TP53, IDH1, BRAF | 55% | ATRX, CTNNB1, PTCH1 | 32% |
| Sarcoma (25) | 40% | TP53, BARD1,<br>CDK12 | 84% | BCOR, H3F3A,<br>IGF2R | 12% |
| Wilms tumor (3) | 34% | RET, TP53 | 67% | DROSHA, TERT | 34% |
| Neuroendocrine (21) | 20% | TP53, ERBB2,<br>NF1 | 39% | SETD2, AKAP9,<br>ANKRD24 | 62% |
| Mesothelioma (5) | 20% | TP53 | 80% | BAP1, BCL2L1,<br>BLNK | 20% |
| Meningioma (235) | 10% | PIK3CA,<br>SMARCB1, ATM | 79% | NF2, AKT1,<br>SMO | 21% |
| Schwannoma (112) | 6% | SMARCB1, ATR,<br>MET | 91% | NF2, AFF3,<br>EMSY | 10% |
| Pituitary adenoma (129) | 4% | ATR, NBN, NRAS | 45% | GNAS, ASPM,<br>ERBB4 | 56% |
| Adrenal (4) | 0% | None | 50% | FLNA, NLRP1,<br>RB1 | 50% |
| Thymoma (1) | 0% | None | 0% | None | 100% |
| Langerhans (1) | 0% | None | 0% | None | 100% |

Conversely, the largest proportion of cancer types in our cohort containing very few mutations in Level 1 genes are meningiomas, which are limited to AKT1 and predominantly exhibit mutations in the Level 2 gene NF2 (**Figure 6**, **Table 3**). Other cancers with few Level 1 variants, or in which more than half of patient samples had no aberration reported, include genomically silent tumors such as pituitary adenomas (68/129) and neuroendocrine cancer (12/21). Actionable mutations in pituitary adenomas are rare, with only 4 out of 129 cases (3.1%) presenting mutations in Level 1 genes, underscoring the need for broader diagnostic NGS approaches in this cancer type.

We also examined the distribution of copy number variation across the ten cancer types with the highest number of CNV-positive cases (**Supplementary Figure 8**). Among cases with clinically reported copy number aberrations (31%, 417/1339), the most reported CNV was NF2 (27%), followed by CDKN2A (20%), EGFR (19%), and PTEN (12%). The most frequent CNV events were hemizygous losses, particularly in meningioma and schwannoma, followed by amplifications in glioblastoma and glioma. Lung cases exhibited a particularly high proportion of amplifications, with 27 of 34 cases affected.

Loss of NF2 — often as part of chromosome 22q loss — was most frequently detected as hemizygous loss (90 cases) in schwannomas and meningiomas, aligning with NF2 frameshift deletions and stop-gain mutations noted above. The majority of tumors containing CNV in NF2 also carried an SNV mutation (97%), leading to biallelic loss of NF2. Following this pattern of biallelic inactivation, alterations in PTEN were predominantly observed in glioblastoma and glioma (84% of combined-loss cases, 31/37). Combined alterations involving APC were primarily found in colorectal, prostate, and lung cancers, while TP53 was reported across a diverse range of malignancies including colorectal, glioblastoma, lung, and pancreatic cancers.

### Clinical Impact of Variants

Commonly identified mutations in specific cancer types include TP53, EGFR, and KRAS variants in lung cancer, and NF2 in schwannomas and meningiomas (**Table 3**, **Figure 7**). Known therapeutic Level 1 amino acid changes in our reported cohort include AKT1 E17K in meningioma and IDH1 R132H (Level 1 in other cancer types), reported in 86% of IDH1-mutated cases, with the remaining arginine 132 IDH1 mutations (R132C, R132G, R132S) reported in 2–6% of cases.

**Figure 7.**
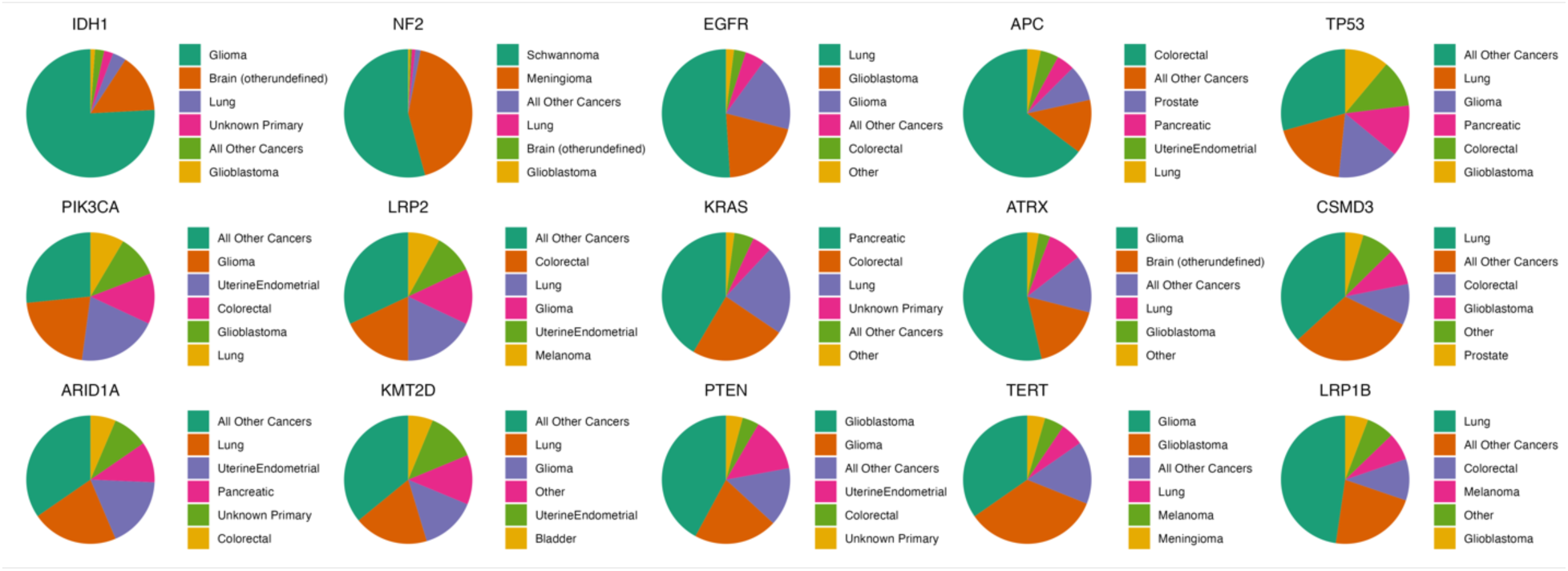
Distribution of cancer types per variant genes. The percent distribution of cancer types associated with the top 15 variant genes by the NYU LG-PACT assay (percentages omitted for clarity, see Supplementary Table 5). For each gene, the five most frequently associated cancer types are shown individually. All remaining cancer types are grouped under the category “All Other Cancers”.

Therapeutic hotspot variants such as BRAF V600E and those in PIK3CA E545 or H1047 are identified broadly across multiple cancer types. BRAF also carries 15 other unique amino acid alterations beyond V600E in our cohort, and 40 additional PIK3CA amino acid substitutions were reported across various cancer types (**Supplementary Table 3**). Of note is the low frequency of breast cancer samples in our cohort, for which therapeutic PIK3CA interventions are FDA approved; only 1 of 5 breast cancer patients was reported with a PIK3CA variant (K111E).

KRAS G12, G13, and Q61 alterations resemble previously reported cancer profiles. The only approved druggable KRAS variant, G12C, is predominantly associated with lung cancer, while other amino acid changes — G12D (investigational), G12R, and G12V — are mainly reported in pancreatic cancer (**Supplementary Figure 9, Supplementary Table 4**). Known oncogenic or likely oncogenic variants in ERBB2 such as p.Y772_A775dup, p.V777L, and p.V842I are also reported. Other genes with a wide range of amino acid substitutions from somatic mutations include NF1, NOTCH1, RB1, MTOR, ALK, and ROS1 **(Supplementary Table 3**), likely complicating efforts to understand and therapeutically target their mechanisms of action.

Additional significant CNV findings include EGFR, MDM4, and PDGFRA locus amplifications seen in GBM, and frequent complete loss of CDKN2A in neurological and pancreatic cases. The arm-scale co-deletion of 1p/19q — a characteristic feature of gliomas — was also identified, though it is currently not reportable by PACT. Oncogenes such as EGFR, CDK4, PDGFRA, and MYC were predominantly reported as amplifications, while tumor suppressor genes NF2, CDKN2A, TP53, and APC were commonly reported as hemizygous losses or deletions (**Supplementary Figure 7**).

Patient samples containing both SNV and CNV occurred in highly heterogeneous cancer types such as glioblastoma and other brain cancers, as well as gastrointestinal cancers in our cohort (**Figure 6**). Notably, all 27 cancer cases designated as “unknown primary” contained at least one reportable variant, with 48% (13/27) having both SNV and CNV. A total of 202 genes were reported across these 27 patients; the most common was TP53 (10 of 27 samples), and 7 (25%) were positive for KRAS G12 mutations, providing valuable genomic information to aid in disease identification and treatment selection. Specifically, one patient sample diagnosed as “metastatic of unknown primary” exhibited an oncogenic profile of SNVs in KRAS, TP53, and APC along with MYC amplification, suggesting a potential colorectal cancer of origin.

## Discussion

Molecular diagnostic applications for cancer patients represent a rapidly evolving field, driven by declining sequencing costs and the continuous discovery of diagnostic, prognostic, and therapeutic biomarkers — advances that have spurred the development of increasingly sophisticated technical assays. Here, we present a comprehensive landscape of genetic mutations identified in a prospective pan-cancer clinical cohort using a 606-gene panel. We further examine how the detection of genetic alterations influences clinical management and demonstrate the practical utility of this approach for treating oncologists.

We applied rigorous clinical thresholds to ensure high-confidence variant calling, incorporating strict quality control criteria, orthogonal assay concordance, false positive elimination, and well-defined limits of detection within our variant reporting protocol and bioinformatic workflow — all of which are publicly available. We describe the genomic findings from sequencing data of the first implementation of our health center’s NYU LG-PACT assay and evaluate clinically reported variants across multiple cancer types. Therapeutic targetability was used as a proxy for clinical utility; encouragingly, the majority of profiled patients harbored at least one variant, and half of those variants occurred in a druggable gene.

As with most targeted sequencing panels, a key limitation of our approach is the reliable identification of small structural variations (indels) and larger structural abnormalities. Only one-third of our cohort had detectable copy number variations, a finding that likely reflects a combination of biological factors — such as cancer type and stage — and technical considerations including tumor content, the sensitivity of variant-calling tools, and the inherently limited genomic breadth of targeted sequencing.

One meaningful measure of clinical utility is the proportion of patients who receive actionable genomic information from a given assay. In our cohort, patients with lung, gastrointestinal, schwannoma, and colorectal cancers derived the greatest benefit from our 606-gene panel, while those with prostate cancer or sarcomas showed comparatively fewer actionable findings. Brain cancers exhibited variable utility: glioblastoma and glioma profiling yielded meaningful treatment guidance, whereas meningioma profiling more commonly provided prognostic or diagnostic value.

By necessity, clinical and pathology laboratories often lag several months to a year or more behind the latest research developments, given the rigorous validation and compliance requirements before any new technology can be applied in a patient care setting. Since the initial approval of our NYU LG-PACT panel, we now additionally have received conditional approval to report tumor mutational burden (TMB) and microsatellite instability (MSI), extending the panel’s clinical scope. Also however at our health institution other essential biomarker testing is still performed either by other NGS platforms (such as for the identification of fusion genes), single gene assays, ddPCR, or companion diagnostics (such as for liquid based screening assays).

Looking ahead, advances in technology demand that we increasingly integrate multiple data modalities into patient management. In tumor board settings, physicians frequently raise critical questions about how a patient’s full genomic profile should inform treatment decisions — how identified variants collectively contribute to the underlying mechanism of disease, and how they can be applied in an individualized, N-of-1 clinical context for the patient. These discussions are essential within our institution and should be broadly shared across the medical community. This includes training medical students in NGS-based genomic diagnostics, publishing case reports that document the application of specific NGS technologies, developing more stringent evidence-based frameworks for variant classification, using agentic AI based tools that collect data and evaluate from past treatment decisions and assessments such as tumor boards, establishing better patient follow-up systems to track and evaluate how real world healthcare data from genomic profiling translates into meaningful outcomes for cancer patients across all institutional settings.

## Data Availability

The datasets generated and/or analyzed during the current study are not publicly available due to Institutional IRB Requirements but are compiled in searchable database format and available from the corresponding author on reasonable request. Pipelines and analysis code is freely available on the NYU-Molecular-Pathology GitHub repository.

## Acknowledgments

We would like to acknowledge significant contributions of Kelsey Zhu (formerly NYU Langone Health Dept. of Molecular Pathology – Bioinformatics) to overall pipeline development, coding of analysis functions and nextflow design.

