## Supplementary Figures for "Genome Profiling of Actionable Cancer Targets (NYU LG-PACT) for Clinical Patient Molecular Diagnostics and Treatment"

### Supplementary Figure 1.

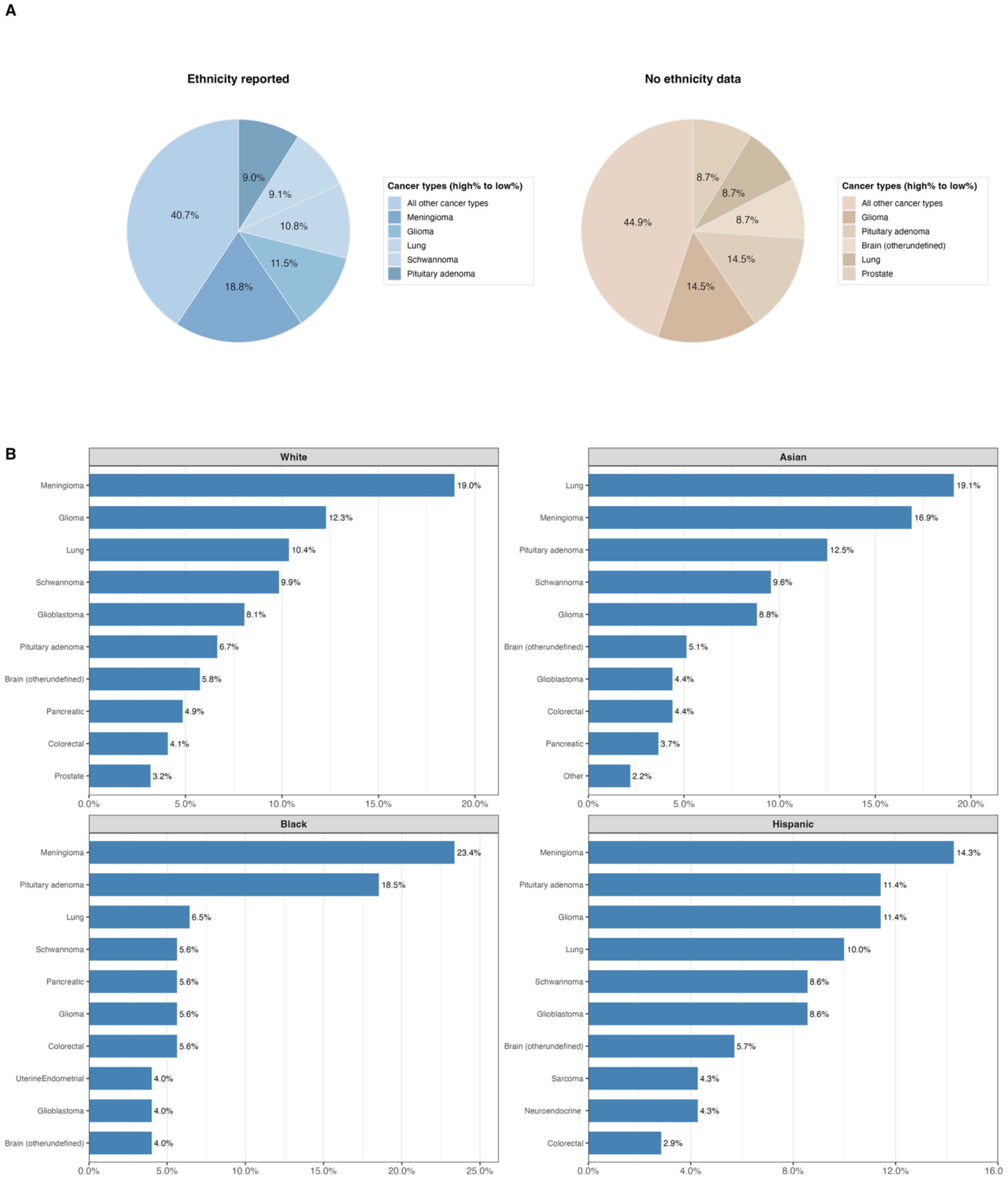


**Supplementary Figure 1. Cancer type distribution by ethnicity reporting and by major ethnicity group.**
**(A)** Proportions of the top five cancer types plus an “All other cancer types” category among cases with ethnicity reported (left) versus those with no ethnicity data (right).

**(B)** For each major ethnicity category (White, Asian, Black, Hispanic), horizontal bars show the top ten most frequent cancer types. Bar length indicates the percentage of cases with that cancer type among all cases assigned to that ethnicity. Cancer types are ordered from most to least frequent within each panel.

#### Supplementary Figure 2.

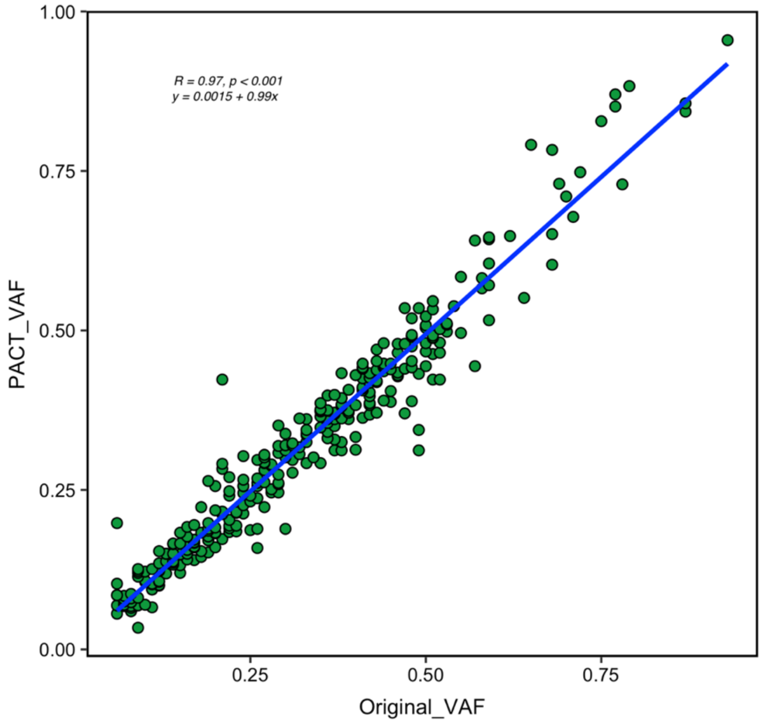


**Supplementary Figure 2. Orthogonal Assay Validation.** NYU LG-PACT Variant Allele Frequency (VAF) was significantly correlated with orthogonally validated variants.

#### Supplementary Figure 3.

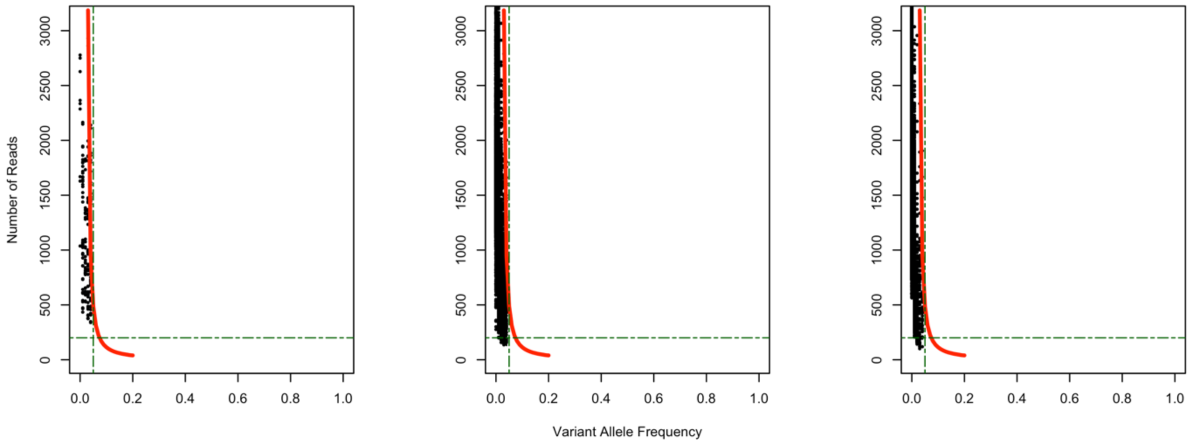


**Supplementary Figure 3. Variant calls below clinically validated thresholds.** All SNVs and Indels as identified empirically with MuTect2 and LoFreqSomatic variant callers below validated thresholds below 5% VAF. Red line – 95% confidence interval (Cheng D., et al. JMDI 2015:17), Sample variants (both SNV and Indels) called by both callers (**A**), MuTect2 (**B**) and LoFreqSomatic (**C**). Green dotted lines – horizontal line represents coverage 200x threshold, vertical line represents VAF thresholds at 5%.

#### Supplementary Figure 4.

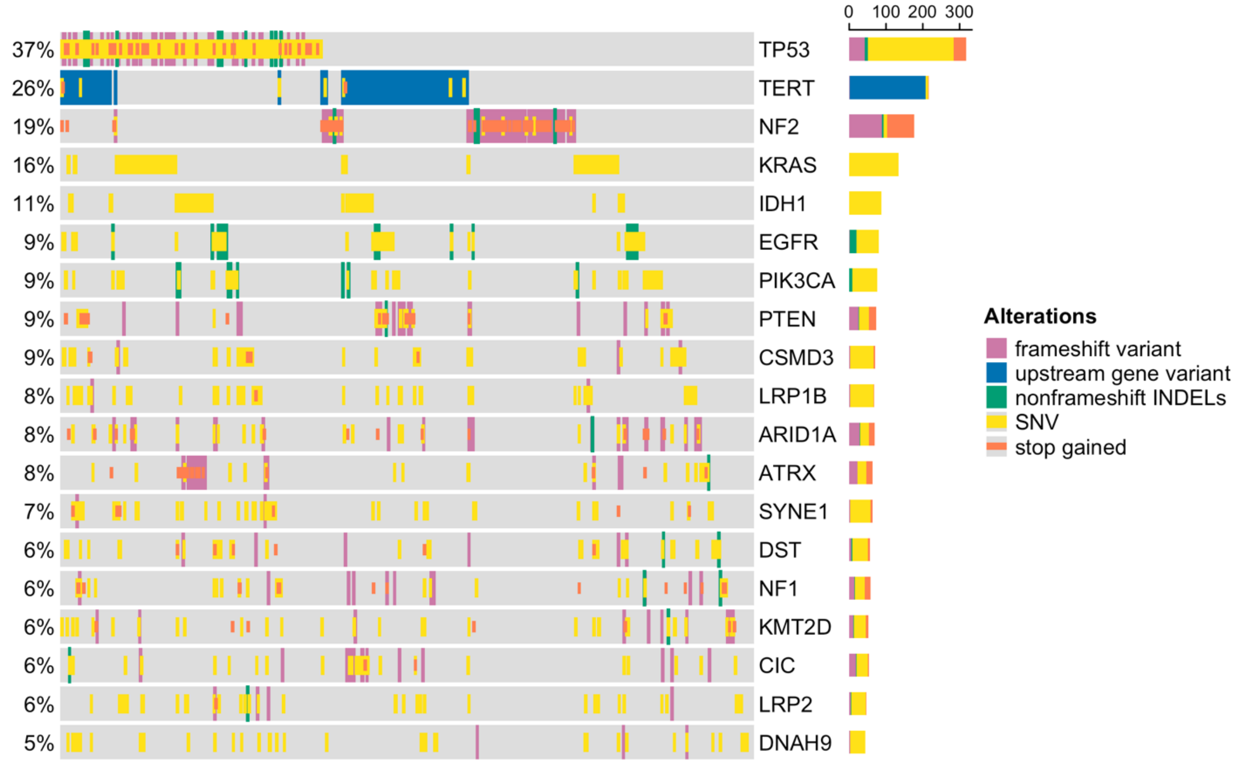


**Supplementary Figure 4. Somatic mutations reported by NYU LG-PACT.** The landscape of somatic mutations for the top 19 pathologist reported genes in our dataset, along with the TERT promoter (profiled only targeted promoter region). Cases included (n=827) only which have at least one reported variant in the top 19 genes or in the TERT promoter. Side histogram depicts count of type of alterations per gene.

#### Supplementary Figure 5.

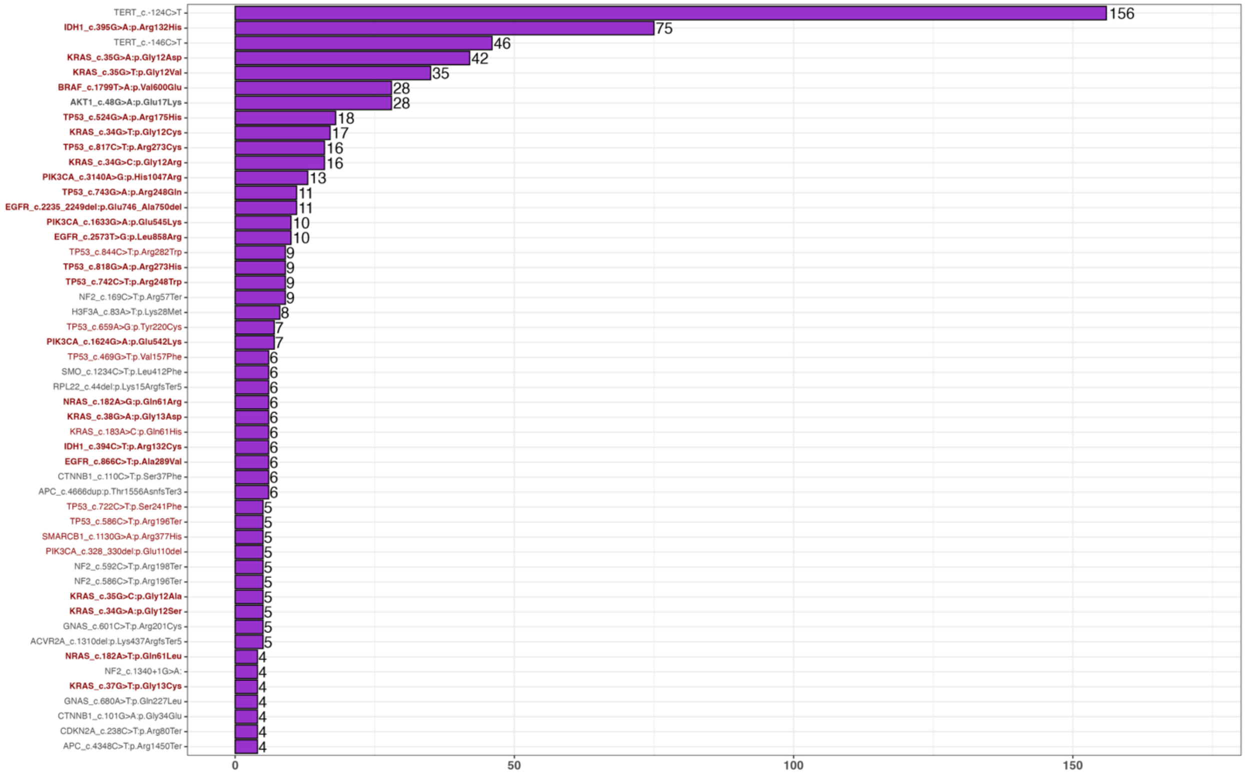


**Supplementary Figure 5. Most Frequent Variants (SNV/Indels) reported by NYU LG-PACT.** Histogram of the top 50 most frequently reported SNV and INDELs by NYU pathologists. Variants are ordered by descending frequency (number of cases reported, x-axis), with counts shown at the end of each bar. On the y-axis, variants occurring within known mutational hotspots are highlighted in bold. Variants leveled in dark red are located in OncoKB$^{TM}$ Level 1 genes. Variants meeting both criteria use labels in both styles.

#### Supplementary Figure 6.

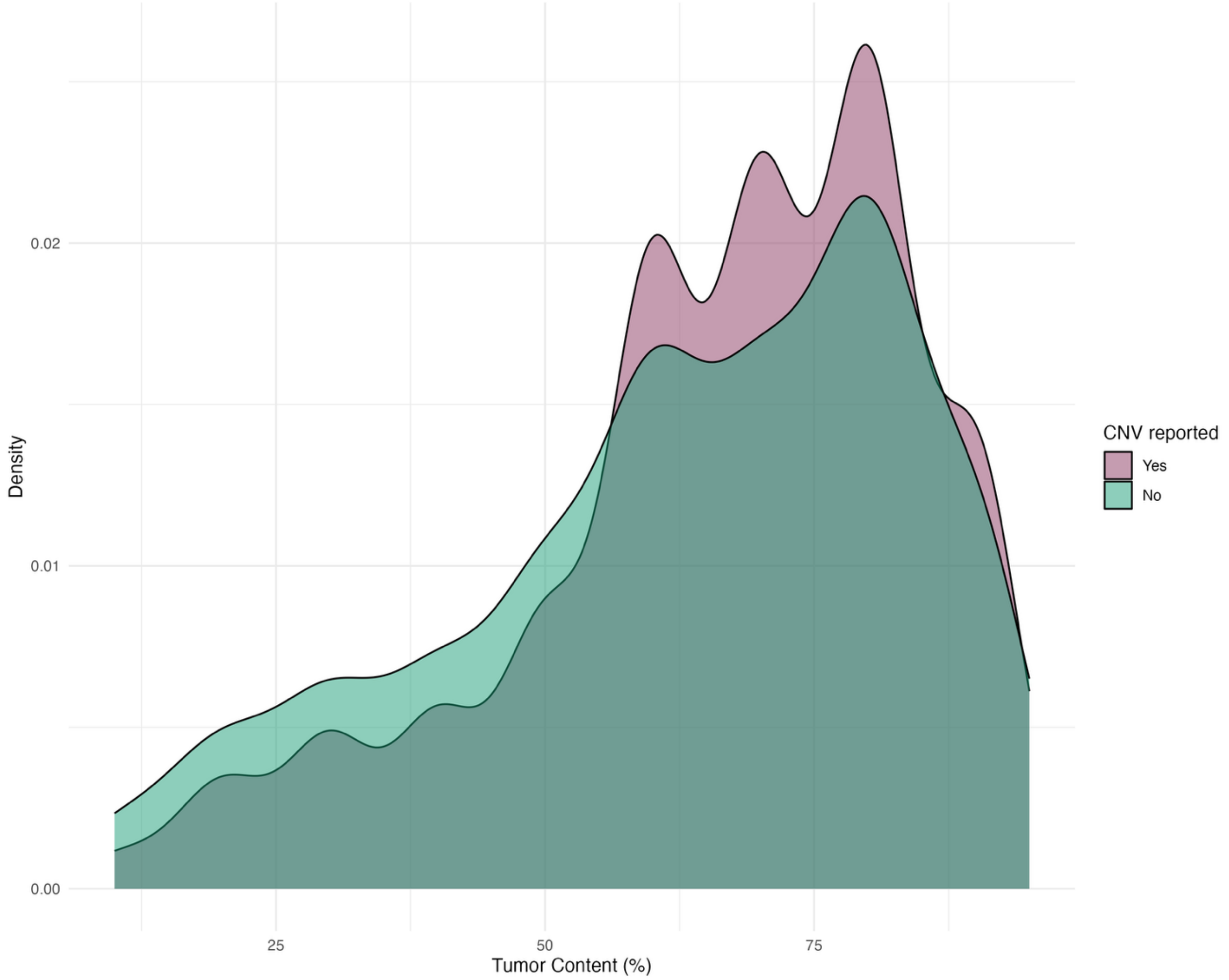


**Supplementary Figure 6. Relationship between tumor content and CNV detection.** Density plot of tumor content (%) across the entire cohort, colored by whether CNVs were reported in the case. The red distribution corresponds to cases with at least one CNV reported, and the green distribution corresponds to cases with no CNVs reported.

#### Supplementary Figure 7.

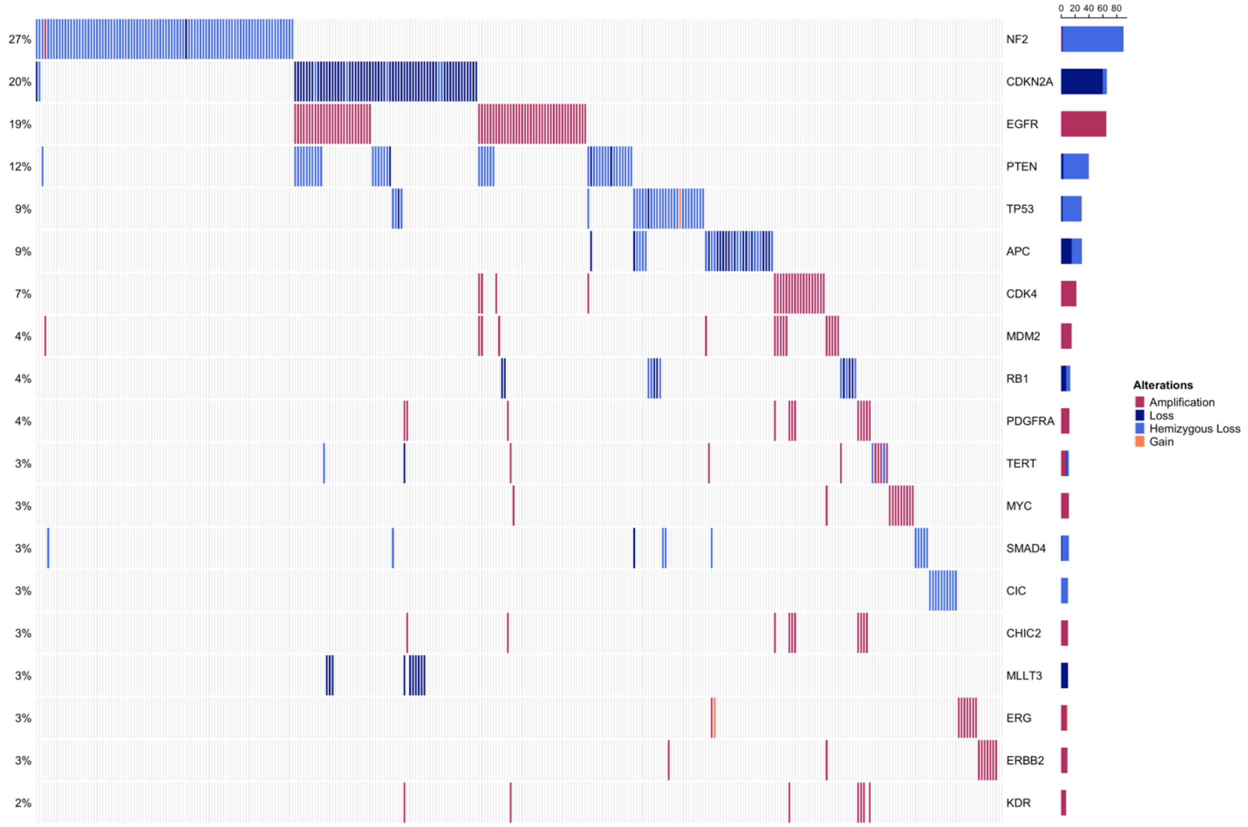


**Supplementary Figure 7. Top CNV genes reported in LG-PACT.** Copy-number oncoplot for the most frequently altered genes among the top–copy-number–reported genes (n = 19). Each column represents one case and each row represents one gene. Color indicates alteration type (amplification, gain, hemizygous loss, loss). The left axis reports the percentage of the cohort with any copy-number event in the selected top 19 genes. The right-hand stacked barplots summarize the count of each alteration class per gene.

#### Supplementary Figure 8.

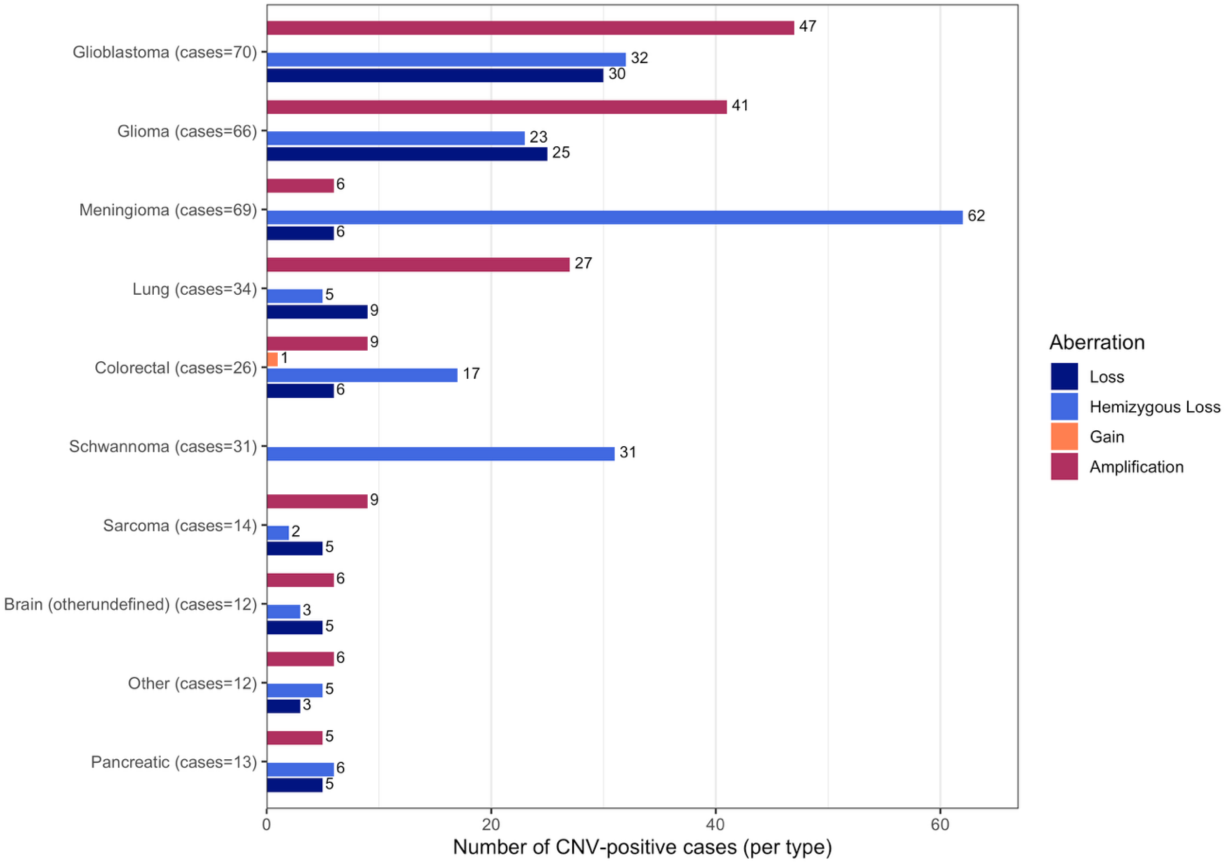


**Supplementary Figure 8. Copy number variation distribution across the top 10 cancer types.** Bar plots showing the number of CNV-positive cases in the 10 cancer types with the highest CNV burden. Bars are stratified by aberration type (loss, hemizygous loss, gain, amplification) and grouped by cancer type. Each bar represents the number of cases with at least one CNV of that aberration type in the indicated cancer, with the case count annotated at the end of the bar.

#### Supplementary Figure 9.

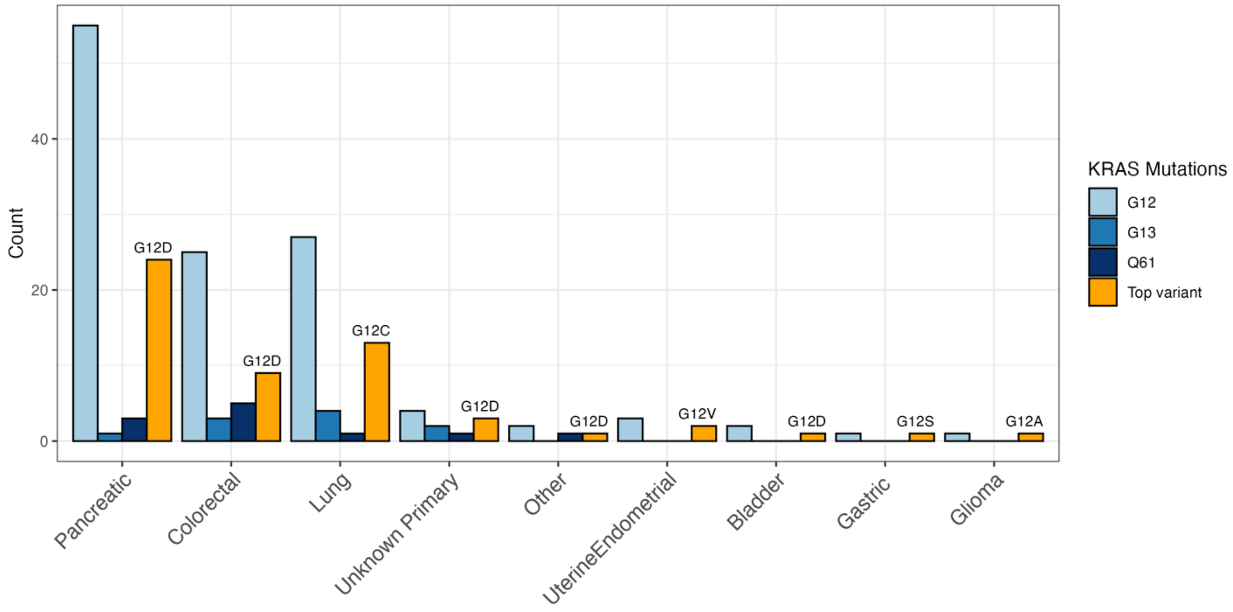


**Supplementary Figure 9**. **KRAS mutations identified by NYU LG-PACT.** Bar plot showing the frequency of KRAS amino acid substitutions per cancer type in the NYU LG-PACT patient cohort. Bars are grouped by cancer types and colored by mutation codon (G12, G13, or Q61). For each cancer type, the right-most (orange) bar highlights the most frequent specific KRAS variant observed (labeled above the bar).
